# COVID-19 and frontline health workers in West Africa: a scoping review

**DOI:** 10.1101/2020.10.15.20213249

**Authors:** Kingsley K. A. Pereko, Edward Kwabena Ameyaw, Shaibu Bukari, Victoria Acquaye, Alfred Dickson Dai-Kosi

**Affiliations:** Department of Community Medicine, School of Medical Sciences, University of Cape Coast, Cape Coast, Ghana; School of Public Health, Faculty of Health, University of Technology Sydney, Australia; University of Cape Coast, Cape Coast, Ghana; Psychological Medicine and Mental Health, School of Medical Sciences, University of Cape Coast, Cape Coast, Ghana; Department of Community Dentistry, School of Medicine and Dentistry, University of Ghana, Legon

**Keywords:** COVID-19, frontline, health workers, healthcare, global health, West Africa

## Abstract

**Introduction:** The novel Coronavirus 2019 (COVID-19) has become a severe global health threat since its emergence. Overcoming the virus is partly dependent on the holistic wellbeing of frontline health workers. Implications of COVID-19 on frontline health workers in West Africa could be substantial given the limited resources and logistics. This scoping review maps available literature on the impact of COVID-19 on frontline health workers in West Africa.

**Materials and methods:** Literature on the impact of COVID-19 on frontline health workers in West Africa were searched in six databases namely Cochrane Library, PubMed, EMBASE, Google Scholar, Africa Journals Online (AJOL) and CINAHL. Further search was done across websites of the ministries of health of West African countries and notable organisations. We conducted a narrative synthesis of the findings taking cognisance of the overarching purpose of the study and the research question.

**Results:** Of the 67 studies identified, 19 were included in the final synthesis. Three main themes emerged and these are impact of COVID-19 on frontline health workers, drivers of susceptibility to COVID-19 and government/donor support. A greater number of the studies originated from Nigeria. Each study reported at least one impact of COVID-19 on frontline health workers in West Africa. The impacts included death, fear, unwillingness to attend to COVID-19 patients and stigmatisation. Some health workers were not adhering to the safety protocols coupled with periodic shortage of personal protective equipment (PPE) and thereby had an increased susceptibility.

**Conclusion:** Being the first scoping review on the impact of COVID-19 on frontline health workers in West Africa, the study has illustrated the urgent need for West African governments to enact laws/rules that would compel all frontline health workers to adhere to all the COVID-19 protocols at the workplace. To end intermittent shortage or issue of inadequate PPEs, governments ought to liaise with local industries by empowering them, providing financial support and creating a conducive atmosphere for them to produce cost effective PPEs using available local resources.

**Scoping review registration:** DOI 10.17605/OSF.IO/B9NXZ (Open Science Framework)

## Introduction

The novel Coronavirus 2019 (COVID-19) has become a severe global health threat since its emergence [1]. As of 5:15pm CEST, 1 October 2020, there were 33,842,281 cases of COVID-19 with 1,010,634 deaths globally [2]. The pandemic has affected both low and middle-income countries as well as high income countries. The role of frontline healthcare providers is therefore indispensable in the combat against COVID-19. These health workers are expected to have a close exposure to COVID-19 infected persons at varying stages of the infection thereby increasing their susceptibility and tendency of further spread [3]. In spite of the fact that COVID-19 has impacted the global community, the virulence level and impact vary across environmental, demographic, socio-economic and demographic spheres [4].

Admittedly, healthcare providers have been impacted by the COVID-19 in high income countries such as the UK and USA [3]. However, the impact for frontline health workers in low and middle income countries such as those in West Africa may be substantial owing to a number of factors. For instance, in addition to the fact that health indices in West African countries are the lowest in the world, the pandemic can easily overwhelm the ailing health systems across West Africa, which are operated by inadequate health personnel [5]. In addition, implications of COVID-19 on frontline health workers in West Africa could be substantial given the limited resources and logistics [6]. All countries in West Africa are in either low or middle income category with little budget allocation to the health sector, thus ranging between 0.6% and 3.4% [5]. The sparse distribution and rural nature of some locations could pose difficulty in sending an infected health worker from a rural setting to a tertiary or secondary level health facilities which are predominantly in urban locations [7]. Consequently, the ability of countries in the sub-region to finance and implement the

requisite measures needed to protect frontline health workers and boost the health systems to rise to the pandemic may be compromised.

Overcoming the novel COVID-19 is partly dependent on the holistic wellbeing of frontline health workers [7]. Even though protection of frontline health workers against COVID-19 is a priority global concern [8, 9], no scoping review have been executed to collate the magnitude of impact on frontline health workers, the specific factors that increase susceptibility and measures instituted to protect frontline health workers from the impacts of the virus in West Africa. COVID-19 has been confirmed in all sixteen (16) West African countries [10]. By September 22^nd^ 2020, a total of 172,594 cases and 2,580 deaths had been recorded in the sub-region [11].

The WHO reports that COVID-19 infections among frontline health workers usually occur at the work place [12]. This underscores the need to explore the situation in West Africa to unravel the documeted impact of the virus on frontline health workers. Previous studies on COVID-19 in West Africa are predominantly reviews focusing on specific countries [13-17]. This is therefore the first scoping review to collate evidence on the impact of COVID-19 on frontline health workers in West Africa guided by the question: “How has COVID-19 impacted frontline healthcare providers in West Africa?” Outcome of the study would not only inform governments and policy makers on the specific socio-culturally sensitive policies required to safeguard the wellbeing of the limited frontline health workers but would as well prompt frontline health workers on what they could do at the personal level to mitigate their susceptibility to the virus whilst taking care of COVID-19 and other patients.

## Materials and methods

We conducted a scoping review between July and October 2020 with respect to the guidelines of the Joanna Briggs Institute’s Preferred Reporting Items for Systematic Reviews and Meta-Analyses Extension for Scoping Reviews (PRISMA-ScR) checklist [18]. This study was guided by a protocol registered with the Open Science Framework (DOI 10.17605/OSF.IO/B9NXZ).

### Population of interest

The review focused on frontline health workers in West Africa. Frontline health worker included any category of healthcare provider who has been providing healthcare and have been directly interacting with patients from 2019 to August 2020.

### Eligibility criteria and study selection

Citations of all published articles were first exported to EndNote and subsequently to the Covidence online systematic review platform. Three key steps were followed to screen the studies; (a) deduplication, (b) title and abstract screening and, (c) full text screening. In the case of the grey literature, selection for inclusion was strictly based on the assessment outcome as well as inclusion and exclusion criteria of the study. Included studies satisfied the following: (i) conducted in West Africa, (ii) reports about frontline health workers, (iii) focuses on any impact of COVID-19, (iv) employed experimental/quasi-experimental, observational, quantitative, qualitative, mixed methods, (v) reports/editorials/commentaries, (vi) in English, French or Spanish (vii) and published between December 2019 and August 2020. Out of the total studies, 19 were included in the final synthesis (see Figure 1).

**Figure 1.**
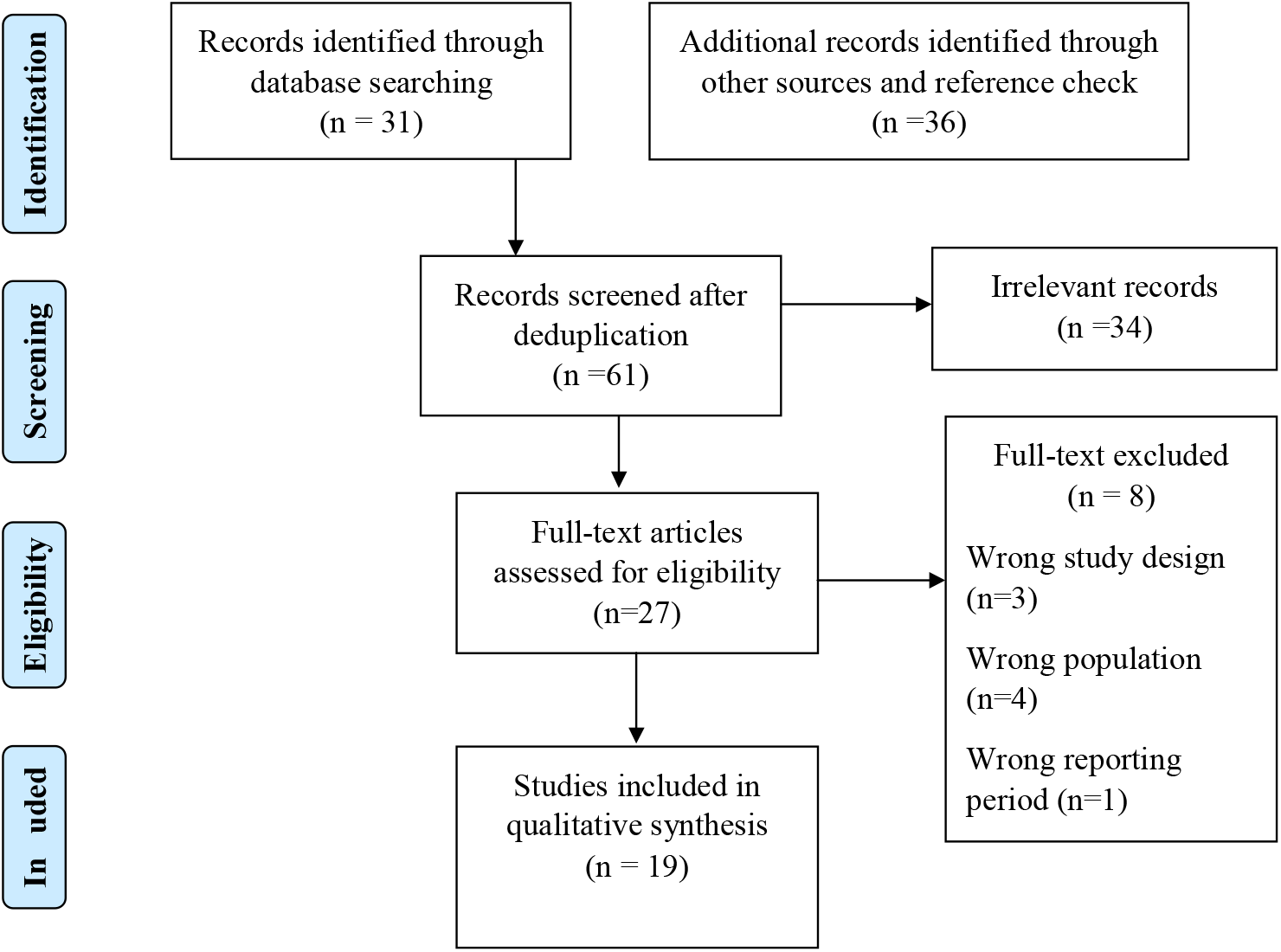
PRISMA 2009 Flow Chart

### Sources of data and search strategy

We conducted an electronic search for both peer-reviewed articles and grey literature. Six databases were searched for published articles: Cochrane Library, PubMed, EMBASE, Google Scholar, Africa Journals Online (AJOL) and CINAHL. We searched for grey literature (e.g. reports, press periodic briefings) from the websites of ministries of health of all the sixteen West African countries and websites of reputable agencies that report on COVID-19 situation in West Africa such as the Africa Centres for Disease Control and Prevention (Africa CDC), and WHO Regional Office for Africa. Our search followed four cardinal steps: (1) use of search terms for articles in the six aforesaid databases; (2) search for grey literature from websites of key organisations and ministries of health of each West African country; (3) manual search for commentaries/editorials/opinions and; (4) manual search of reference lists of included article. A complete search strategy and key words used for PubMed have been provided (S1).

### Quality assessment

The quality assessment was conducted independently by one author (EKA) and verified by another author (KAP). This was done with the McMaster Critical review [19]. The Authority, Accuracy, Coverage, Objectivity, Date, and Significance (AACODS) Checklist was used to assess the quality of the non-peer reviewed studies [20].

### Data charting

We developed a data charting form and this was used to extract important data required to address the overarching review question. The form constituted the following components: (a) author(s) and year of publication, (b) country/scope of study, (c) title, (d) study type, (e) study design, (f) theme and; (g) key findings.

### Synthesis and reporting of findings

We carried out a thematic analysis of the findings from the included studies. Thematic analysis is “a method for identifying, analyzing and reporting patterns within data.” [21]. Three principal themes emerged and the findings were categorised according to these themes. Subsequently, we interpreted and conducted a narrative synthesis of the findings taking cognisance of the overarching purpose of the study and the research question.

## Results

### Characteristics of included studies/reports

A total of 19 studies were included in the synthesis and these are summarised in Table 1. Most of the studies were reviews (n=7) and there was only one original research article which used a quantitative approach [22]. All studies were conducted in 2020, in English and most were from Nigeria (n=8).

**Table 1.** Characteristics of included studies/reports.

| # | Author (year) | Country/Scope | Title | Study type | Study design |
| --- | --- | --- | --- | --- | --- |
| 1. | Ogolodom et al. (2020) | Nigeria | Knowledge, Attitudes and Fears of HealthCare Workers towards the Corona Virus Disease (COVID-19) Pandemic in South-South, Nigeria | Original research article | Quantitative |
| 2. | UNICEF (2020) | Guinea-Bissau | GUINEA-BISSAU: COVID-19 Situation Report – #17 | Report | n/a |
| 3. | Darboe M.K. (2020) | Gambia | Gambia's health system near collapse amid pandemic | Personal reflection | n/a |
| 4. | Shaban A.R.A.<br>(2020) | Ghana | Ghana coronavirus:<br>29,672 cases; 100-bed<br>specialist hospital ready | News article | n/a |
| 5. | Nakri E.<br>(2020) | Ghana | Striving to keep health<br>worker infections at bay | Personal<br>reflection | n/a |
| 6. | Quakyi N.K.<br>(2020) | Ghana | Ghana's much praised<br>COVID-19 strategy has<br>gone awry. Here is why | Commentary | n/a |
| 7. | Agence de<br>Presse<br>Africaine<br>(APA) (2020) | Ghana | Ghana: Press highlights<br>recovery of 1870 health<br>workers from Covid-19,<br>others | Review | n/a |
| 8. | CGTN Africa<br>(2020) | Guinea-Bissau | Nearly 10% of Guinea-Bissau health workers<br>infected with COVID-19 | Review | n/a |
| 9. | Brown W.<br>(2020) | Guinea-Bissau | Hospitals in tiny<br>Guinea-Bissau<br>'overwhelmed' by the<br>pandemic | Review | n/a |
| 10 | WHO (2020a) | Africa | COVID-19 WHO<br>African Region External<br>Situation Report 15 | Report | n/a |
| 11. | Karmo H. | Liberia | Liberia: Minister of | News article | n/a |
|  | (2020) |  | Health Attributes<br>COVID-19 Infections<br>among Health Workers<br>to ‘State of Denial’ of<br>Nurses |  |  |
| 12. | Tih F. (2020) | Nigeria | Nigeria: 800 health<br>workers infected with<br>COVID-19 | Personal<br>reflection | n/a |
| 13. | Okunola<br>A.(2020) | Nigeria | 5 Challenges Facing<br>Health Care Workers in<br>Nigeria as They Tackle<br>COVID-19 | Review | n/a |
| 14. | Clottey P. &<br>Dauda M.<br>(2020) | Nigeria | Striking Doctors in<br>Nigeria Demand<br>COVID-19 PPE, Hazard<br>Pay | Review | n/a |
| 15. | Nwosu-Igbo N<br>(2020) | Nigeria | In the frontline of<br>Nigeria’s struggle with<br>COVID-19 | Review | n/a |
| 16. | Onyeji E.<br>(2020) | Nigeria | COVID-19: As more<br>health workers get<br>infected, JOHESU<br>distributes PPEs | News article | n/a |
| 17. | Amnesty<br>International | Nigeria | Nigeria: Authorities<br>must protect health | Review | n/a |
|  | (2020) |  | workers on the frontline<br>of COVID-19 response |  |  |
| 18. | Mwai P. &<br>Giles C.<br>(2020) | Nigeria | Coronavirus: How<br>vulnerable are health<br>workers in Nigeria? | News article | n/a |
| 19. | WHO (2020b) | Africa | COVID-19 WHO<br>African Region External<br>Situation Report 23 | Report | n/a |

### Narrative synthesis

Three principal themes emerged from the included studies: (a) impact of COVID-19 on frontline health workers; (b) drivers of susceptibility to COVID-19 and; (c) government/donor support. All the records associated with each theme and sub-theme are summarised in Table 2.

**Table 2:** Themes and sub-themes from included studies.

| Theme | Key findings | Included studies |
| --- | --- | --- |
| Impact of COVID-19 on frontline health workers | Infected frontline health workers (n=14) | [13, 14, 16, 17, 23-32] |
|  | Fear of being at risk<br>(n=3) | [17, 22, 28] |
|  | Reduced willingness to<br>go to work (n=1) | [22] |
|  | Worry (n=1) | [34] |
|  | Sadness (n=1) | [34] |
|  | Death (n=2) | [28, 33] |
|  | Stigmatization (n=1) | [29] |
|  | Mental health (n=1) | [29] |
|  | Separation from<br>families (n=1) | [29] |
| Drivers of susceptibility |  |  |
|  | Insecure workplace<br>environment (n=1) | [22] |
|  | Non-adherence to safety<br>protocols/carelessness<br>(n=2) | [26, 34] |
|  | Inadequate<br>infrastructure/equipment<br>(n=7) | [13, 15-17, 28, 33, 35] |
| Government/Donor<br>support (e.g. UNICEF) | Training (n=2) | [23, 35] |
|  | Equipment acquisition | [14, 16, 35] |
|  | /distribution (n=3) |  |
|  | Physical infrastructure<br>(n= 1) | [24] |
|  | Social services (n=1) | [35] |

#### Theme 1: impact of COVID-19 on frontline health workers

Almost all included studies reported at least one impact of COVID-19 on frontline health workers. The dominant impact was COVID-19 infection among frontline health workers such as doctors and nurses as reported by fourteen studies [13, 14, 16, 17, 23-32]. Some health care providers expressed fear of being at risk of contracting COVID-19 and subsequent death [17, 22, 28, 33] as well as expression of worry and sadness [34]. COVID-19 had also reduced the passion or willingness to work in Nigeria [22], and also brought about stigmatisation and separation from families [29].

#### Theme 2: drivers of susceptibility to COVID-19

Three dominant factors were noted to increase frontline health workers’ susceptibility to COVID-19 in West Africa. The commonly reported was inadequate infrastructure/equipment predominantly from Nigeria [15, 16, 28, 33, 35], Ghana [17] and Guinea-Bissau [13]. In the case of Ghana [34] and Liberia [26], non-adherence to the COVID-19 safety protocols among frontline health workers was reported whilst insecure work environment [22] was recounted as factor that enhances susceptibility of frontline health workers to COVID-19 in Nigeria [22].

#### Theme 3: Government/Donor support

To alleviate the impact of COVID-19 on frontline health workers, governments have adopted varied interventions and strategies. A number of non-governmental and international organisations have also supported. For instance, the United Nations International Children’s Emergency Fund (UNICEF) appeared to have assisted in diverse ways. Specific approaches for combating the COVID-19 among frontline healthcare providers include training as evidenced in Guinea-Bissau and Nigeria [23, 35] as well as acquisition and distribution of personal protective and other essential equipment as reported from The Gambia [14], and Nigeria [16, 35]. The support have also manifested in physical infrastructure [24] and provision of social services (e.g. encouraging preventive actions in communities through risk communications) [35] in Ghana and Nigeria respectively.

## Discussion

This review is the first to synthesise evidence on the impact of COVID-19 on frontline health workers in West Africa. The review has illustrated the peculiar implications of COVID-19 on frontline health workers in West Africa, factors that increase their susceptibility and ongoing support/commitment by governments and donor organisations.

A key theme from the review is that COVID-19 has affected and continues to affect frontline health workers in diverse ways with infection among frontline health workers emerging as the dominant impact. Others were death, fear of being at risk, worry, attenuated preparedness to work, stigmatisation and insecure workplace. This indicates that West Africa contributes to the globally estimated 30,000 deaths among frontline health workers [36]. Due to these implications, some health workers are reluctant to attend to COVID-19 patients even if adequately compensated [22]. Similar reports have emerged from other countries outside West Africa such as Mexico, Saudi Arabia and Pakistan where death, worry and mental health issues were noted among frontline health workers [37-39]. The findings indicate the need for extra care and support for West African based frontline health workers especially during pandemics because the overwhelmed health systems further present challenging times for health workers [40]. To this end, putting in place sustainable insurance policy [22], ensuring safe, decent work conditions and intermittent psychological services for frontline health workers may be required to mitigate these implications in West Africa [40]. These can be achieved through inter-sectoral collaboration between governments, employers and workers’ organisations. More importantly, stakeholders’ ability to contexualise protective measures in line with local resources and inter-country nuances might be prudent.

The review identified three factors that incline frontline health workers to COVID-19 in West Africa; insecure workplace environment, non-adherence to COVID-19 safety protocols or carelessness and inadequate infrastructure and equipment including Personal Protective Equipment (PPE). These reflect both systemic gaps and negligence on the part of frontline health workers. In as much as West African governments and their partners are obliged to ensure safety and holistic wellbeing of frontline health workers amidst the COVID-19, the frontline health workers also have an essential role to play in order to ameliorate the situation. Some level of discipline is required by the frontline health workers to ensure their own safety because non-use of PPEs or non-adherence of the COVID-19 protocols is as perilous government’s refusal to purchase and distribute such lifesaving resources or equipment.

More workshops on COVID-19 protocols, constant reminders (e.g. through text messaging, audio-visuals) and sanctioning of frontline health workers who ignore the protocols may help to ensure that the all frontline health workers in West Africa are committed to ensuring their own safety. Further, frontline health workers who are sensitive to the COVID-19 protocols and those who fully observe the protocols may be incentivised to motivate others to do same. Government and partner organisations, however, may have to intervene to ensure safe workplace for the frontline health workers by expanding infrastructure and ensuring consistent supply of PPEs. Regular training of health workers in emergency preparedness can make them conscious and responsive the pandemic and subsequent disease outbreaks.

The study revealed that the support offered to frontline health workers by West African governments and donor organisations manifest in training, acquisition and distribution of equipment, physical infrastructure and social services. These illustrate that governments of various West African countries and some donor partners have instituted some measures with respect to infrastructure and equipment with the aim of boosting the health systems to overcome the novel COVID-19. In Gambia, for instance, the government has expended $12 million on equipment to support the country’s health by acquiring ventilators, ambulances and PPEs since March 2020 to support the overwhelmed health system and expedite the combat against COVID-19 [14]. A number of West African countries such as Ghana and Nigeria have done same through the assistance of partner organisations such as the WHO [41, 42]. West African countries can also establish health emergency funds to cushion the health systems during disease outbreaks.

However, our evidence support previous findings on the inadequacy and intermittent shortage of essential PPEs among frontline health workers [43, 44]. Most of these PPEs are imported [45] and could possibly account for the intermittent shortages. It is therefore imperative for West African countries to ulitise local resources to develop domestic PPEs whenever possible in order not to be over reliant on international trade. This is essentially critical considering that the pandemic compelled most countries to close their borders, a situation that do not permit international transfer of PPEs.

## Strengths and limitations

Most of the included studies were not peer-reviewed. This is due to the recency of COVID-19 and its late entry into West Africa compared to other sub-regions. The review focused on only frontline health workers and hence could not account for the impact of COVID-19 on other category of health workers within West Africa.

## Conclusion

Being the first scoping review on the impact of COVID-19 on frontline health workers in West Africa, the review has highlighted the specific impacts, as well as essential systemic and health personnel gaps reinforcing the impact. The review has also revealed ongoing support and commitment by governments and partner organisations. There is an urgent need for West African governments to enact laws/rules that would compel all frontline health workers to adhere to all the COVID-19 protocols at the workplace. Effective supervision may be essential for achieving full implementation of such laws/rules. To end intermittent shortage or issue of inadequate PPEs, governments ought to liaise with local industries by empowering them, providing financial support and creating a conducive atmosphere for them to produce cost effective PPEs using available local resources. More empirical studies are required to better understand the country specific and contextual factors associated with the impact of COVID-19 on frontline health workers across the sixteen West African countries.

## Supporting information

PRISMA-ScR-Checklist

## Data Availability

All data are included in the manuscript.

## Acknowledgements

None

## PRISMA 2009 Checklist

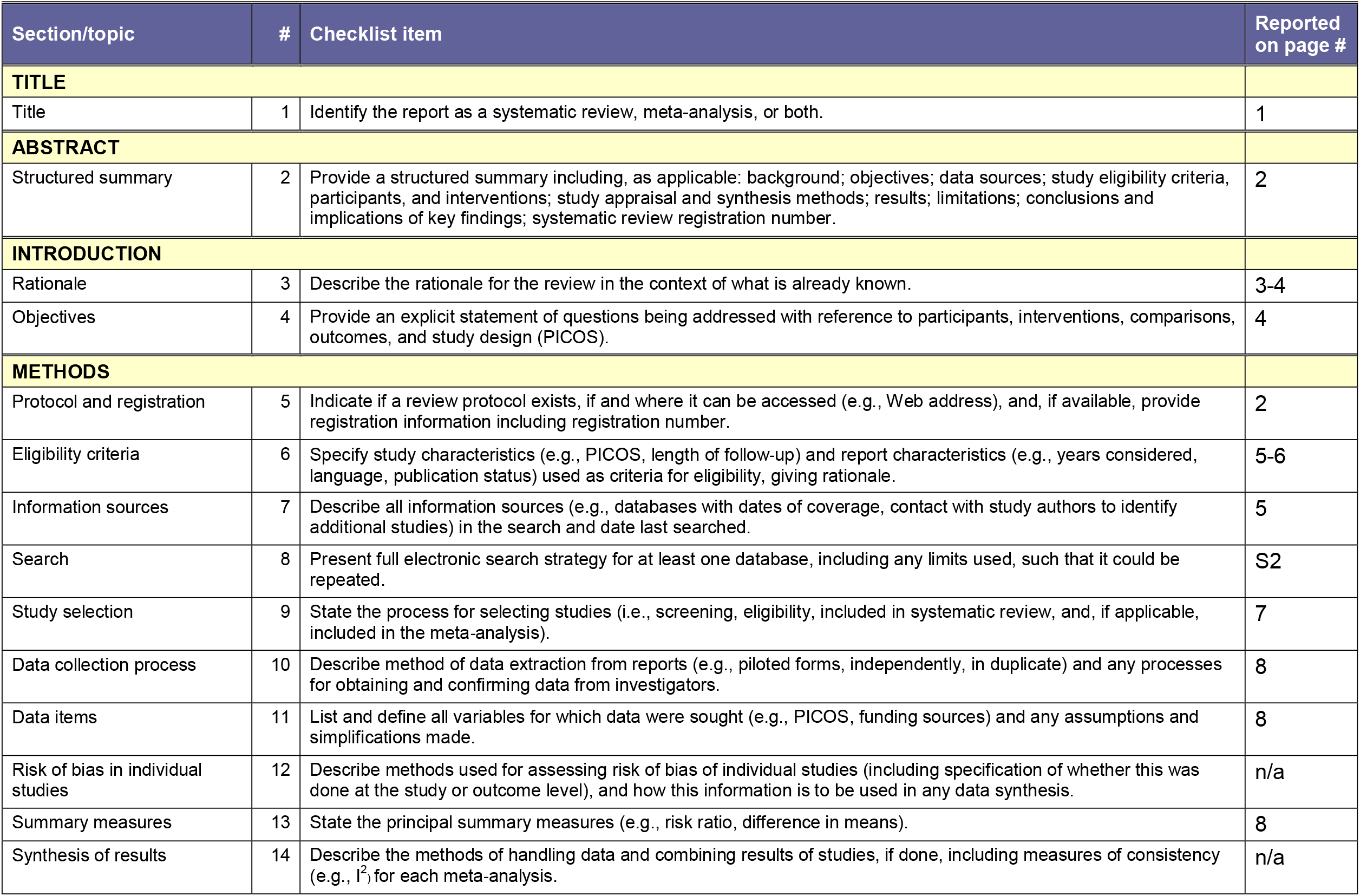
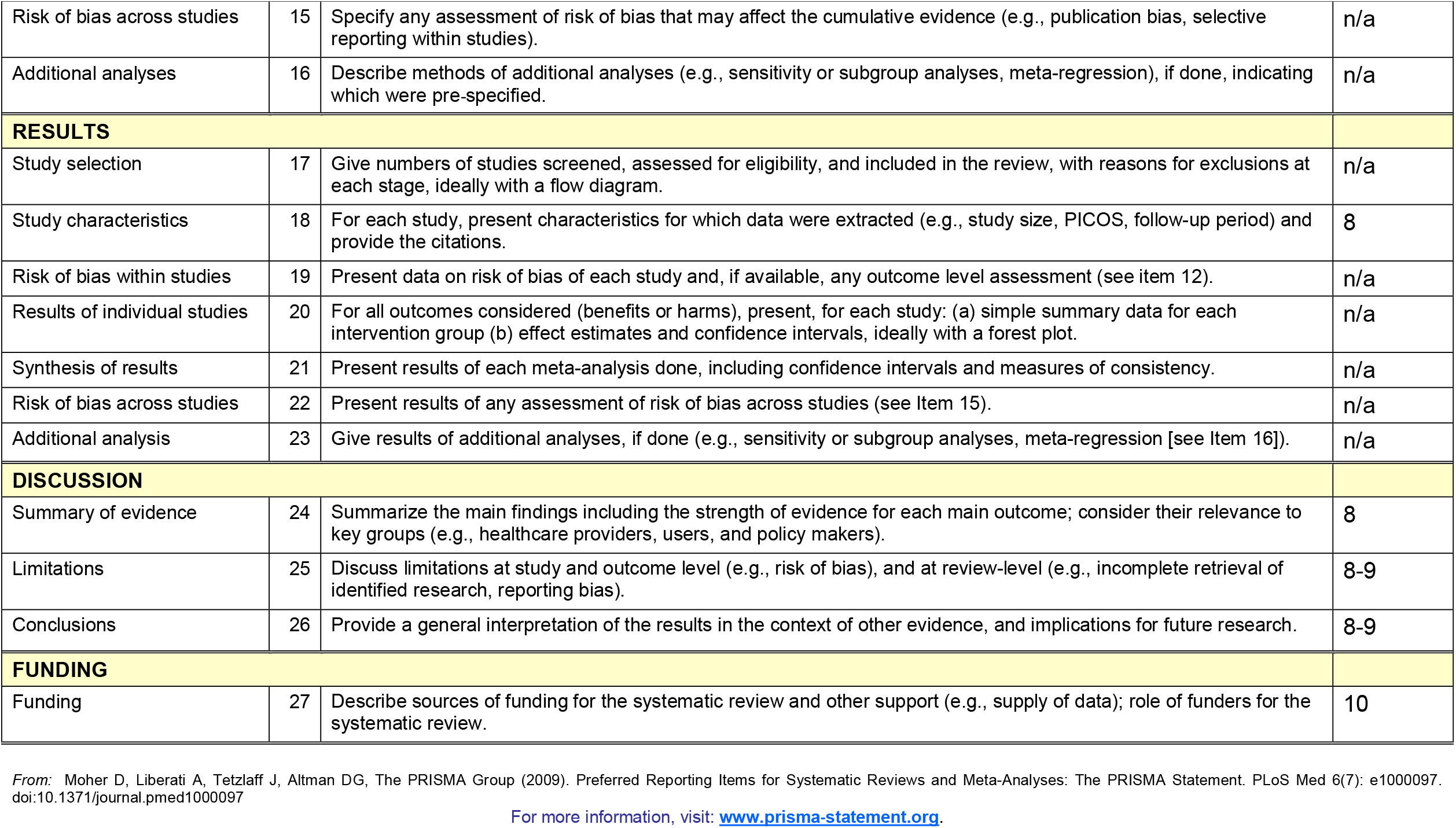

| Section/topic | # | Checklist item | Reported on page # |
| --- | --- | --- | --- |
| **TITLE** |  |  |  |
| Title | 1 | Identify the report as a systematic review, meta-analysis, or both. | 1 |
| **ABSTRACT** |  |  |  |
| Structured summary | 2 | Provide a structured summary including, as applicable: background; objectives; data sources; study eligibility criteria, participants, and interventions; study appraisal and synthesis methods; results; limitations; conclusions and implications of key findings; systematic review registration number. | 2 |
| **INTRODUCTION** |  |  |  |
| Rationale | 3 | Describe the rationale for the review in the context of what is already known. | 3-4 |
| Objectives | 4 | Provide an explicit statement of questions being addressed with reference to participants, interventions, comparisons, outcomes, and study design (PICOS). | 4 |
| **METHODS** |  |  |  |
| Protocol and registration | 5 | Indicate if a review protocol exists, if and where it can be accessed (e.g., Web address), and, if available, provide registration information including registration number. | 2 |
| Eligibility criteria | 6 | Specify study characteristics (e.g., PICOS, length of follow-up) and report characteristics (e.g., years considered, language, publication status) used as criteria for eligibility, giving rationale. | 5-6 |
| Information sources | 7 | Describe all information sources (e.g., databases with dates of coverage, contact with study authors to identify additional studies) in the search and date last searched. | 5 |
| Search | 8 | Present full electronic search strategy for at least one database, including any limits used, such that it could be repeated. | S2 |
| Study selection | 9 | State the process for selecting studies (i.e., screening, eligibility, included in systematic review, and, if applicable, included in the meta-analysis). | 7 |
| Data collection process | 10 | Describe method of data extraction from reports (e.g., piloted forms, independently, in duplicate) and any processes for obtaining and confirming data from investigators. | 8 |
| Data items | 11 | List and define all variables for which data were sought (e.g., PICOS, funding sources) and any assumptions and simplifications made. | 8 |
| Risk of bias in individual studies | 12 | Describe methods used for assessing risk of bias of individual studies (including specification of whether this was done at the study or outcome level), and how this information is to be used in any data synthesis. | n/a |
| Summary measures | 13 | State the principal summary measures (e.g., risk ratio, difference in means). | 8 |
| Synthesis of results | 14 | Describe the methods of handling data and combining results of studies, if done, including measures of consistency (e.g., $I^2$) for each meta-analysis. | n/a |
| Risk of bias across studies | 15 | Specify any assessment of risk of bias that may affect the cumulative evidence (e.g., publication bias, selective reporting within studies). | n/a |
| Additional analyses | 16 | Describe methods of additional analyses (e.g., sensitivity or subgroup analyses, meta-regression), if done, indicating which were pre-specified. | n/a |
| **RESULTS** |  |  |  |
| Study selection | 17 | Give numbers of studies screened, assessed for eligibility, and included in the review, with reasons for exclusions at each stage, ideally with a flow diagram. | n/a |
| Study characteristics | 18 | For each study, present characteristics for which data were extracted (e.g., study size, PICOS, follow-up period) and provide the citations. | 8 |
| Risk of bias within studies | 19 | Present data on risk of bias of each study and, if available, any outcome level assessment (see item 12). | n/a |
| Results of individual studies | 20 | For all outcomes considered (benefits or harms), present, for each study: (a) simple summary data for each intervention group (b) effect estimates and confidence intervals, ideally with a forest plot. | n/a |
| Synthesis of results | 21 | Present results of each meta-analysis done, including confidence intervals and measures of consistency. | n/a |
| Risk of bias across studies | 22 | Present results of any assessment of risk of bias across studies (see Item 15). | n/a |
| Additional analysis | 23 | Give results of additional analyses, if done (e.g., sensitivity or subgroup analyses, meta-regression [see Item 16]). | n/a |
| **DISCUSSION** |  |  |  |
| Summary of evidence | 24 | Summarize the main findings including the strength of evidence for each main outcome; consider their relevance to key groups (e.g., healthcare providers, users, and policy makers). | 8 |
| Limitations | 25 | Discuss limitations at study and outcome level (e.g., risk of bias), and at review-level (e.g., incomplete retrieval of identified research, reporting bias). | 8-9 |
| Conclusions | 26 | Provide a general interpretation of the results in the context of other evidence, and implications for future research. | 8-9 |
| **FUNDING** |  |  |  |
| Funding | 27 | Describe sources of funding for the systematic review and other support (e.g., supply of data); role of funders for the systematic review. | 10 |

From: Moher D, Liberati A, Tetzlaff J, Altman DG, The PRISMA Group (2009). Preferred Reporting Items for Systematic Reviews and Meta-Analyses: The PRISMA Statement. PLoS Med 6(7): e1000097. doi:10.1371/journal.pmed1000097
For more information, visit: www.prisma-statement.org.

