## Supplementary material for "COVID-19 and frontline health workers in West Africa: a scoping review": PRISMA-ScR-Checklist

**Preferred Reporting Items for Systematic reviews and Meta-Analyses extension for Scoping Reviews (PRISMA-ScR) Checklist**

|  | **SECTION** |  |  | **ITEM** |  |  | **PRISMA-ScR CHECKLIST ITEM** |  |  | **REPORTED** |
| --- | --- | --- | --- | --- | --- | --- | --- | --- | --- | --- |
|  |  |  |  |  |  |  |  |  |  | **ON PAGE #** |
|  | **TITLE** | |  |  |  |  |  |  |  |  |
|  | Title | | 1 | |  |  | Identify the report as a scoping review. | |  | 1 |
|  | **ABSTRACT** | |  |  |  |  |  |  |  |  |
|  |  |  |  |  |  |  | Provide a structured summary that includes (as | |  |  |
|  | Structured | |  |  |  |  | applicable): background, objectives, eligibility criteria, | |  |  |
|  |  |  | 2 | |  |  | sources of evidence, charting methods, results, and | |  |  |
|  | summary | |  |  |  |  |  |  |  | 2 |
|  |  |  |  |  |  |  | conclusions that relate to the review questions and | |  |  |
|  |  |  |  |  |  |  | objectives. | |  |  |
|  | **INTRODUCTION** | |  |  |  |  |  |  |  |  |
|  |  |  |  |  |  |  | Describe the rationale for the review in the context of | |  |  |
|  | Rationale | | 3 | |  |  | what is already known. Explain why the review | |  |  |
|  |  |  |  |  |  |  | questions/objectives lend themselves to a scoping | |  |  |
|  |  |  |  |  |  |  |  |  |  | 4-5 |
|  |  |  |  |  |  |  | review approach. | |  |  |
|  |  |  |  |  |  |  | Provide an explicit statement of the questions and | |  |  |
|  |  |  |  |  |  |  | objectives being addressed with reference to their key | |  |  |
|  | Objectives | | 4 | |  |  | elements (e.g., population or participants, concepts, and | |  | 5 |
|  |  |  |  |  |  |  | context) or other relevant key elements used to | |  |  |
|  |  |  |  |  |  |  | conceptualize the review questions and/or objectives. | |  |  |
|  | **METHODS** | |  |  |  |  |  |  |  |  |
|  |  |  |  |  |  |  | Indicate whether a review protocol exists; state if and | |  |  |
|  | Protocol and | | 5 | |  |  | where it can be accessed (e.g., a Web address); and if | |  |  |
|  | registration | |  |  |  |  | available, provide registration information, including the | |  | 6 |
|  |  |  |  |  |  |  | registration number. | |  |  |
|  |  |  |  |  |  |  | Specify characteristics of the sources of evidence used | |  |  |
|  | Eligibility criteria | | 6 | |  |  | as eligibility criteria (e.g., years considered, language, | |  | 6 |
|  |  |  |  |  |  |  | and publication status), and provide a rationale. | |  |  |
|  |  |  |  |  |  |  | Describe all information sources in the search (e.g., | |  |  |
|  | Information | | 7 | |  |  | databases with dates of coverage and contact with | |  |  |
|  | sources* | |  |  |  |  | authors to identify additional sources), as well as the | |  | 6-7 |
|  |  |  |  |  |  |  | date the most recent search was executed. | |  |  |
|  |  |  |  |  |  |  | Present the full electronic search strategy for at least 1 | |  |  |
|  | Search | | 8 | |  |  | database, including any limits used, such that it could be | |  | 7 |
|  |  |  |  |  |  |  | repeated. | |  |  |
|  | Selection of | |  |  |  |  | State the process for selecting sources of evidence (i.e., | |  |  |
|  | sources of | | 9 | |  |  |  |  |  |  |
|  |  |  |  |  |  |  | screening and eligibility) included in the scoping review. | |  | 7 |
|  | evidence† | |  |  |  |  |  |  |  |  |
|  |  |  |  |  |  |  | Describe the methods of charting data from the included | |  |  |
|  |  |  |  |  |  |  | sources of evidence (e.g., calibrated forms or forms that | |  |  |
|  | Data charting | | 10 | |  |  | have been tested by the team before their use, and | |  |  |
|  | process‡ | |  |  |  |  | whether data charting was done independently or in | |  | 8 |
|  |  |  |  |  |  |  | duplicate) and any processes for obtaining and | |  |  |
|  |  |  |  |  |  |  | confirming data from investigators. | |  |  |
|  | Data items | | 11 | |  |  | List and define all variables for which data were sought | |  |  |
|  |  |  |  |  |  |  | and any assumptions and simplifications made. | |  | 9 |
|  | Critical appraisal of | |  |  |  |  | If done, provide a rationale for conducting a critical | |  |  |
|  |  |  |  |  |  |  | appraisal of included sources of evidence; describe the | |  |  |
|  | individual sources | | 12 | |  |  |  |  |  | 8 |
|  |  |  |  |  |  |  | methods used and how this information was used in any | |  |  |
|  | of evidence§ | |  |  |  |  |  |  |  |  |
|  |  |  |  |  |  |  | data synthesis (if appropriate). | |  |  |
|  | Synthesis of results | | 13 | |  |  | Describe the methods of handling and summarizing the | |  |  |
|  |  |  |  |  |  |  | data that were charted. | |  | 8 |

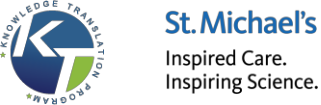

|  | **SECTION** |  |  | **ITEM** |  |  | **PRISMA-ScR CHECKLIST ITEM** |  |  | **REPORTED** |
| --- | --- | --- | --- | --- | --- | --- | --- | --- | --- | --- |
|  |  |  |  |  |  |  |  |  |  | **ON PAGE #** |
|  | **RESULTS** | |  |  |  |  |  |  |  |  |
|  | Selection of | |  |  |  |  | Give numbers of sources of evidence screened, | |  |  |
|  |  |  |  |  |  |  | assessed for eligibility, and included in the review, with | |  |  |
|  | sources of | | 14 | |  |  |  |  |  |  |
|  |  |  |  |  |  |  | reasons for exclusions at each stage, ideally using a flow | |  | 9 |
|  | evidence | |  |  |  |  |  |  |  |  |
|  |  |  |  |  |  |  | diagram. | |  |  |
|  | Characteristics of | |  |  |  |  | For each source of evidence, present characteristics for | |  |  |
|  | sources of | | 15 | |  |  |  |  |  |  |
|  |  |  |  |  |  |  | which data were charted and provide the citations. | |  | 9-12 |
|  | evidence | |  |  |  |  |  |  |  |  |
|  | Critical appraisal | |  |  |  |  | If done, present data on critical appraisal of included | |  |  |
|  | within sources of | | 16 | |  |  |  |  |  | - |
|  |  |  |  |  |  |  | sources of evidence (see item 12). | |  |  |
|  | evidence | |  |  |  |  |  |  |  |  |
|  | Results of | |  |  |  |  | For each included source of evidence, present the | |  |  |
|  | individual sources | | 17 | |  |  | relevant data that were charted that relate to the review | |  | 9-12 |
|  | of evidence | |  |  |  |  | questions and objectives. | |  |  |
|  | Synthesis of results | | 18 | |  |  | Summarize and/or present the charting results as they | |  | 9-14 |
|  |  |  |  |  |  |  | relate to the review questions and objectives. | |  |  |
|  | **DISCUSSION** | |  |  |  |  |  |  |  |  |
|  |  |  |  |  |  |  | Summarize the main results (including an overview of | |  |  |
|  | Summary of | | 19 | |  |  | concepts, themes, and types of evidence available), link | |  | 15-17 |
|  | evidence | |  |  |  |  | to the review questions and objectives, and consider the | |  |  |
|  |  |  |  |  |  |  | relevance to key groups. | |  |  |
|  | Limitations | | 20 | |  |  | Discuss the limitations of the scoping review process. | |  | 17 |
|  |  |  |  |  |  |  | Provide a general interpretation of the results with | |  |  |
|  | Conclusions | | 21 | |  |  | respect to the review questions and objectives, as well | |  | 18 |
|  |  |  |  |  |  |  | as potential implications and/or next steps. | |  |  |
|  | **FUNDING** | |  |  |  |  |  |  |  |  |
|  |  |  |  |  |  |  | Describe sources of funding for the included sources of | |  |  |
|  | Funding | | 22 | |  |  | evidence, as well as sources of funding for the scoping | |  |  |
|  |  |  |  |  |  |  | review. Describe the role of the funders of the scoping | |  | 18 |
|  |  |  |  |  |  |  | review. | |  |  |

JBI = Joanna Briggs Institute; PRISMA-ScR = Preferred Reporting Items for Systematic reviews and Meta-Analyses extension for Scoping Reviews.

- Where *sources of evidence* (see second footnote) are compiled from, such as bibliographic databases, social media platforms, and Web sites.

† A more inclusive/heterogeneous term used to account for the different types of evidence or data sources (e.g., quantitative and/or qualitative research, expert opinion, and policy documents) that may be eligible in a scoping review as opposed to only studies. This is not to be confused with *information sources* (see first footnote).

‡ The frameworks by Arksey and O’Malley (6) and Levac and colleagues (7) and the JBI guidance (4, 5) refer to the process of data extraction in a scoping review as data charting*.*

§ The process of systematically examining research evidence to assess its validity, results, and relevance before using it to inform a decision. This term is used for items 12 and 19 instead of "risk of bias" (which is more applicable to systematic reviews of interventions) to include and acknowledge the various sources of evidence that may be used in a scoping review (e.g., quantitative and/or qualitative research, expert opinion, and policy document).

*From:* Tricco AC, Lillie E, Zarin W, O'Brien KK, Colquhoun H, Levac D, et al. PRISMA Extension for Scoping Reviews

(PRISMAScR): Checklist and Explanation. Ann Intern Med. 2018;169:467–473. [doi: 10.7326/M18-0850.](http://annals.org/aim/fullarticle/2700389/prisma-extension-scoping-reviews-prisma-scr-checklist-explanation)

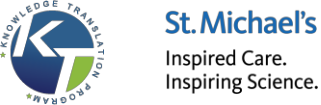

2
